# Building a Prediction Model to Detect Elevated Risk for Developing a Substance Use Disorder

**DOI:** 10.64898/2026.10.16.26365759

**Authors:** I-Tzu Hung, Fazil Aliev, Sally I-Chun Kuo, Alyssa Kim, Geoffrey Khan, Holly E. Poore, Genevieve F. Dash, Maia Choi, Hang Zhou, Antti Latvala, Sari Aaltonen, Jaakko Kaprio, Jessica E. Salvatore, Richard J. Rose, COGA Collaborators, Danielle M. Dick

**Author notes:** Correspondence to Danielle M. Dick PhD: Department of Psychiatry Robert Wood Johnson Medical School Rutgers University Piscataway, NJ 08854.

## Abstract

**<u>Importance</u>:** Preventing substance use disorder (SUD) is a top public health priority, but clinical practice currently lacks validated prognostic tools to identify risk before the onset of heavy use/problems.

**<u>Objective</u>:** To determine whether models can predict SUD using genetic, behavioral, and environmental factors and to evaluate performance across substance types (alcohol, cannabis, tobacco), sex, and genetic similarity groups.

**<u>Design</u>:** This prognostic study developed and validated a multi-domain risk model using data from four longitudinal cohorts (The National Longitudinal Study of Adolescent to Adult Health, Avon Longitudinal Study of Parents and Children, The Collaborative Study on the Genetics of Alcoholism, and FinnTwin12) with follow-up from early adolescence into young adulthood.

**<u>Setting</u>:** Population-based and family-based samples from the United States, United Kingdom, and Finland.

**<u>Participants</u>:** The analysis included participants with available genomic and behavioral data. The cohorts represented a spectrum of risk, ranging from nationally representative community samples to high-risk populations.

**<u>Exposure(s) (for observational studies)</u>:** Predictors included polygenic scores for substance use-related phenotypes and 10 established early-life behavioral and environmental risk factors.

**<u>Main Outcome(s) and Measure(s)</u>:** The outcomes were a diagnosis of moderate-to-severe SUD based on the Diagnostic and Statistical Manual of Mental Disorders, Fifth Edition (DSM-5) criteria. Model performance was evaluated using the area under the receiver operating characteristic curve (AUC), sensitivity, specificity, and positive and negative predictive value.

**<u>Results</u>:** A total of 16,362 participants were included in the study (54.7% female; mean [SD] age 26.73 [3.36]). Genetic similarity group composition was 70.4% European, 16.8% African, 9.8% admixed American, 2.2% East Asian, and 0.7% South Asian. SUD prevalence was 22.0%. The integrated model achieved an AUC of 0.78 for predicting any SUD, with a sensitivity of 69% and a positive predictive value of 45%. Performance was comparable for specific substances (AUC range 0.74–0.82) and across genetic similarity groups (AUC range 0.61-0.82). The model demonstrated higher AUC in females compared with males.

**<u>Conclusions and Relevance</u>:** Findings highlight the potential utility of an SUD prediction model to inform clinicians and patients about risk for developing a SUD.

****Key points**:** *<u>Question</u>:* Can we develop prediction models to detect elevated risk for developing a substance use disorder (SUD) prior to onset.?

*<u>Findings</u>:* Using a harmonized dataset of four diverse longitudinal cohorts, we developed and evaluated a prediction model that integrates genetic, behavioral, and environmental predictors. The model demonstrated good discriminatory power to identify individuals at risk for future SUD.

*<u>Meaning</u>:* These findings establish a framework for integrating multi-domain, multi-informant data into clinical workflows, offering a basis for personalized risk stratification and prevention of SUDs.

## Introduction

Substance use disorders (SUDs) are associated with substantial personal, economic, and societal costs,^1^ with over 48 million individuals in the United States meeting criteria for a SUD.^2^ Preventing substance use problems has been cited as a top public health priority by the US Preventive Services Task Force for over two decades;^3,4^ however, screening remains rare and inconsistent in health care settings.^5–7^ Prevention efforts are also suboptimal in that they are designed to screen for current risky substance use and associated problems (e.g., SBIRT^8^). Ideally, prevention efforts would be designed to avert problems before they occur.

SUDs are prime candidates for early risk prediction and prevention because they are developmental in nature, characterized by a trajectory of risk behavior that emerges long before the onset of substance use problems.^9,10^ Decades of longitudinal research have identified early risk factors that robustly and consistently predict substance use problems, including behavioral and emotional challenges, family history, trauma, and peer characteristics.^11–17^ In addition, genetic studies have made substantial progress in identifying the genetic variants involved in SUDs, with polygenic scores (PGS), calculated by aggregating risk variants across the genome identified via genome-wide association studies (GWAS),^18^ now accounting for 7-10% of the variance in SUDs.^19,20^ Importantly, these studies have also revealed that etiological factors are largely shared across SUDs,^19,21–23^ meaning that the same genetic, behavioral, and environmental factors broadly impact all forms of substance problems, with only a small portion being drug specific. Initial efforts by our team to develop multivariable models that incorporate a broad range of genetic, behavioral, and environmental factors found that individuals in the top decile of combined risk have 4-8 times increased risk of developing a SUD compared to those in the bottom decile.^24^

In this study, we^24^ combine data from four longitudinal cohorts, collected across three countries, and representing both population-based and high-risk samples, to develop prediction models for SUD. Our primary goal was to evaluate prediction and model performance, to lay the foundation for clinical application. Secondary analyses evaluated whether model performance varied for individual substances, across sex, and across genetic similarity groups.

## Methods

### Sample

Four samples were used for model development:

*The National Longitudinal Study of Adolescent to Adult Health (Add Health)*: a nationally representative longitudinal study of US adolescents followed into adulthood.^25^ Data were analyzed from Wave I (1994-1995) when respondents were between 12–19 years, Wave II (1996; ages 13-19) and Wave IV (2008-2009; ages 24-32).

*Avon Longitudinal Study of Parents and Children (ALSPAC)*: an ongoing, longitudinal population-based study of a birth cohort in Southwest England.^26–28^ Pregnant female residents with an expected date of delivery between April 1, 1991 and December 31, 1992 were invited to participate. The initial number of pregnancies enrolled was 14,541, with 13,988 children alive at 1 year of age. This analysis uses data up to the age 24 assessment. Since 2014, study data have been collected and managed using REDCap electronic data capture tools hosted at the University of Bristol.^29^ The ALSPAC study website contains details of all available data through a fully searchable data dictionary and variable search tool https://www.bristol.ac.uk/alspac/researchers/our-data/. Ethical approval for the study was obtained from the ALSPAC Law and Ethics committee and local research ethics committees (NHS Haydock REC: 10/H1010/70).

*Collaborative Study on the Genetics of Alcoholism (COGA)*: a multi-site, multi-generational, family-based sample consisting of alcohol dependent individuals identified through 7 treatment centers across the United States, their extended families, and community controls.^30^ We use the prospective sample of offspring of COGA participants, with baseline assessments at ages 12– 22 and biennial follow-up.^31^

*FinnTwin12 (FT12)*: a population-based study of Finnish twins born 1983–1987 identified through Finland’s Central Population Registry.^32^ Twins completed survey assessments at ages 12, 14, 17, and 22.

Table 1 shows the Ns and substance use descriptives for each sample. We restricted the analytic sample to those with available genotypic, phenotypic, behavioral, and environmental data. Genetic similarity groups were assigned using the Phase 3 reference panel from the 1000 Genomes Project. Principal components and SNP weights were estimated with EIGENSOFT SmartPCA^33^ in the reference data and projected onto the study sample for classification.

**Table 1.** Sample Characteristics by Cohort.

|  | Add Health |  |  |  |  | ALSPAC | COGA |  | FT12 |
| --- | --- | --- | --- | --- | --- | --- | --- | --- | --- |
| Genetic similarity group | EUR | AFR | AMR | EAS | SAS | EUR | EUR | AFR | EUR |
| Sample size<sup>a</sup> | 5050 | 1845 | 1608 | 366 | 112 | 3673 | 1748 | 910 | 1050 |
| Female | 53.5% | 55.6% | 50.2% | 44.5% | 53.6% | 61.1% | 51.5% | 52.9% | 53.5% |
| Any SUD | 29.9% | 13.6% | 20.5% | 17.8% | 25.9% | 7.5% | 39.9% | 34.4% | 15.1% |
| AUD | 17.9% | 5.6% | 11.3% | 9.8% | 15.2% | 5.4% | 27.5% | 15.8% | 13.3% |
| TUD | 10.3% | 3.1% | 4.6% | 2.7% | 7.1% | 1.4% | 8.2% | 4.5% | 1.9% |
| CUD | 7.0% | 6.5% | 7.5% | 5.2% | 8.9% | 0.6% | 22.8% | 28.5% | 0.9% |

Notes: Any SUD, AUD, TUD, and CUD were defined as meeting criteria for moderate to severe SUD. AFR=African-like; AMR=Admixed American-like; EAS=East Asian-like; EUR=European-like; SAS=South Asian-like, as defined by genomic data.
<sup>a</sup>Available samples with genotypic, phenotypic, and environmental risk data.

### Measures

#### SUD diagnosis

The primary outcome was meeting criteria for any SUD. We leveraged all available substance use assessments across each of the four cohorts to create harmonized diagnoses, aligning with DSM-5 criteria whenever possible. Alcohol use disorder (AUD), tobacco use disorder (TUD), and cannabis use disorder (CUD) were assessed in all cohorts; COGA and FT12 additionally assessed opioid, stimulant, sedative, hallucinogen, inhalant, and cocaine use disorders. Details about measures for each cohort are available in eMethods 1 in the Supplement. Cases were defined as meeting criteria for moderate to severe SUD (≥4 symptoms) for any assessed SUD. For TUD, all cohorts administered the Fagerström Test for Nicotine Dependence (FTND)^34^, and a cutoff of ≥7 was used to define cases. Individuals who met the criteria for any of the assessed substances were coded as cases for the any SUD outcome; individuals not meeting the criteria were classified as controls. Included among the controls were individuals who reported 1 or 2 SUD symptoms, since prior research suggests the 2-symptom threshold for mild SUD often captures non-pathological use patterns, raising concerns about over-diagnosis.^35^ We conducted sensitivity analyses comparing model development using SUD outcomes defined by meeting mild-or-above criteria (eMethods 2). Secondary analyses focused on the most commonly used substances: AUD, TUD and CUD.

#### Behavioral and Environmental Risk Index (BERI)

Ten early-life risk factors were included based on their associations with SUD outcomes in the literature: low childhood socioeconomic status (SES), family history of SUD, early initiation of substance use, childhood internalizing problems, childhood externalizing problems, drinking in adolescence, smoking in adolescence, cannabis use in adolescence, peer substance use, and exposure to trauma/stressful experiences. These measures were operationalized identically to those defined in Barr et al.^24^, with the exception that we additionally included a measure of stressful experiences for FT12, defined as endorsement of experiencing a serious illness or injury or the death of a close individual. Each BERI item was dichotomized as present or absent and entered independently as a predictor in the model. Correlations among BERI items were low to moderate (eFigure 1 in the Supplement).

**Figure 1.**
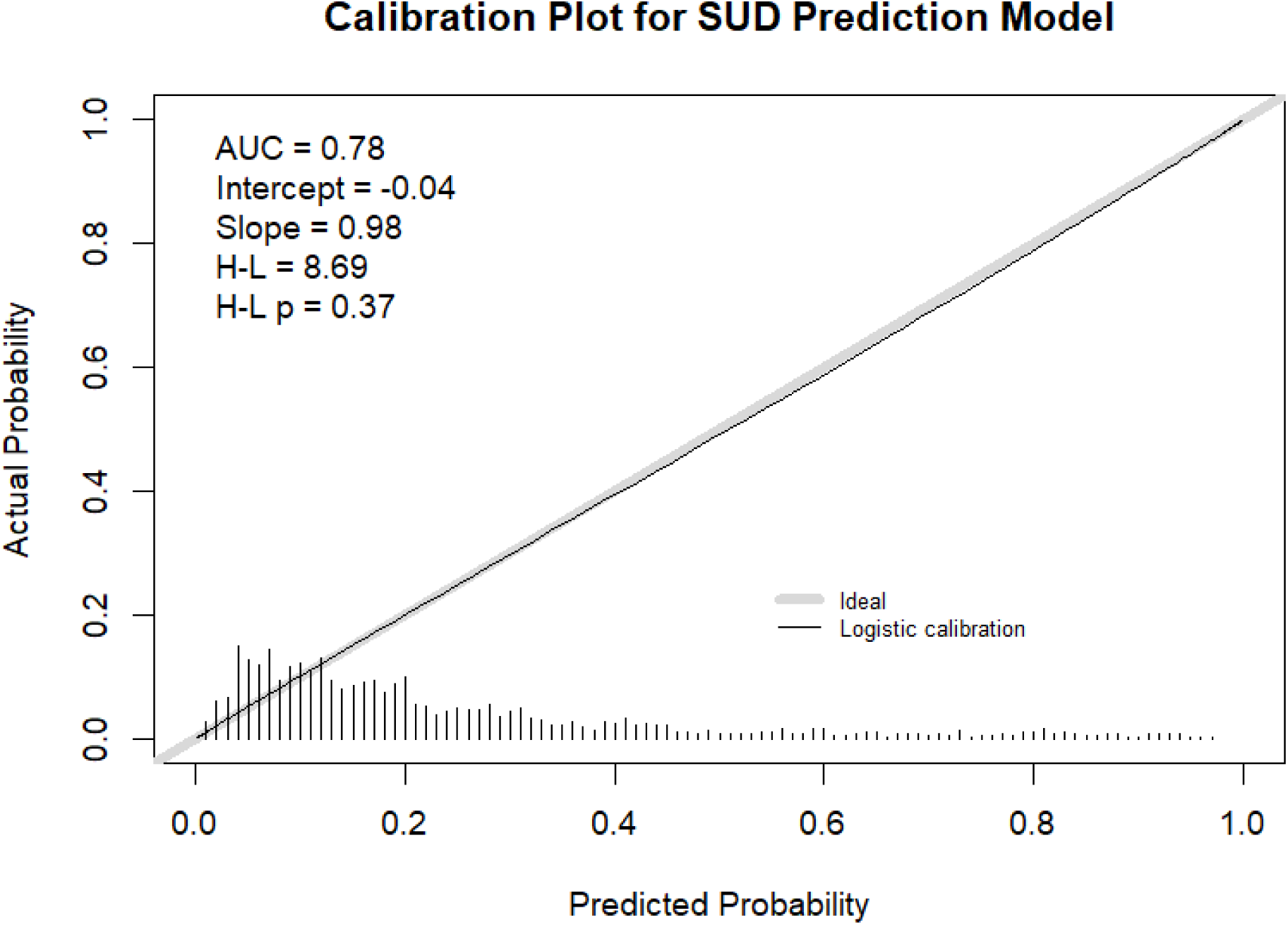
Calibration Plot for SUD Prediction Model Note. H-L = Hosmer-Lemeshow test; H-L p= p value for the Hosmer–Lemeshow test.

#### Genetic Risk

We included several PGS in our models, including an index of liability toward disinhibitory disorders (Externalizing; EXT), as well as substance-specific PGSs for problematic alcohol use (PAU-specific), problematic tobacco use (PTU-specific), cannabis use disorders (CUD-specific), and opioid use disorders (OUD-specific). The latter represent genetic influences specific to substance use outcomes after accounting for the broad genetic liability that impacts all substance use/disinhibitory disorders.^19^ These PGS were calculated from a multivariate GWAS analysis of >2.2 million individuals^19^, with the resulting PGS accounting for the greatest variances in SUDs to date. In addition, a PGS representing liability toward internalizing disorders (INT), including depression, anxiety, PTSD, and neuroticism, was calculated using GWAS summary statistics from the largest discovery sample to date (N > 2.6 million).^36^ Hold-out GWAS were run when any of the 4 cohorts included in the present analyses were part of the original GWAS discovery sample to remove any redundancy with the present sample (eMethods 3 in the Supplement).

PGSs were calculated separately for each genetic similarity group (e.g., European-like and African-like) using weights derived from PRS-CS^37^ and scores generated using PLINK2^38^. To account for population stratification, we followed the pooling approach described in Barr et al.,^24^ regressing PGSs on linear and quadratic effects of age and the first 10 genetic principal components within each genetic similarity group and cohort and standardize the residualized PGSs. These standardized scores were then pooled across the genetic similarity groups and used in subsequent analyses. By doing this, we retained sample size and power while developing one model that can be applied across individuals from various genetic similarity groups.

#### Analytic strategy

To develop and evaluate a model that predicts SUD risk from combined genetic, behavioral, and environmental predictors, we pooled data across cohorts, trained the model on a subset of participants, and then tested its performance on a held-out subset, independent from training. The four cohorts were pooled and harmonized via a fixed-effects integrative data analytic (IDA) approach.^39^ We split each cohort into training (80%) and testing/validation (20%) sets using stratified sampling to maintain balance in key characteristics (e.g., SUD prevalence, sex, genetic similarity group). To prevent data leakage, family members were assigned exclusively to either the training or testing set, ensuring that no family was split across both. Additionally, to account for familial clustering in COGA and FT12, clustering-robust standard error was incorporated. Regression-based models were iteratively developed and refined using 10-fold cross-validation within the training dataset.^40^ Penalized regression models, least absolute shrinkage and selection operator (LASSO) and ridges, were also evaluated but did not improve model performance compared with traditional regression (see eTable 1 in the Supplement). The model validation phase involved applying the trained model to the independent hold-out testing dataset and evaluating model performance. Model discriminatory power was evaluated using the area under the receiver operating characteristic curve (AUC) by plotting sensitivity against 1–specificity. Classification performance was evaluated at three probability thresholds: the optimal threshold (maximizing sensitivity and specificity), the 80th percentile (top 20% of predicted risk), and the 95th percentile (top 5% of predicted risk). Performance at each threshold was assessed using sensitivity, specificity, and positive and negative predictive values (PPV and NPV). In addition, odds ratios (ORs) and relative risks (RRs) comparing individuals above vs below each threshold were calculated. Calibration was assessed via calibration plots and Hosmer-Lemeshow tests (H-L) to evaluate the agreement between predicted risks and observed event rates.

The SUD model was subsequently applied to predict AUD, TUD, and CUD separately to test whether the model performed comparatively for common forms of SUD. Model discrimination, indexed by the AUC, was compared between the general SUD model and each substance-specific model using DeLong’s test.^41^ To evaluate whether the prediction models performed comparably across sex, we applied models for each outcome separately in males and females and compared AUC values. When significant differences were observed, we included sex-by-predictor interaction terms to examine whether they explained additional variance in the outcome and improved classification performance. SUD model performance metrics were also calculated separately for the five genetic similarity groups to assess whether performance differed across groups. Our analytic strategy was preregistered on the Open Science Framework (https://osf.io/7dmq8).

## Results

### Model Performance for General SUD Risk

Model performance metrics are displayed in Table 2. The model AUC was 0.78 for SUD. At the optimal threshold, sensitivity was 0.69 and specificity was 0.75. The PPV (0.45) was approximately twofold higher than the sample prevalence (22%). Higher thresholds (80th and 95th percentiles) were associated with lower sensitivity and NPV but higher specificity and PPV. Threshold-based stratification differentiated high- and low-risk groups. At the optimal threshold, the OR was 6.45 and RR was 4.01. At the 95th percentile threshold, the OR increased to 19.56.

**Table 2.**
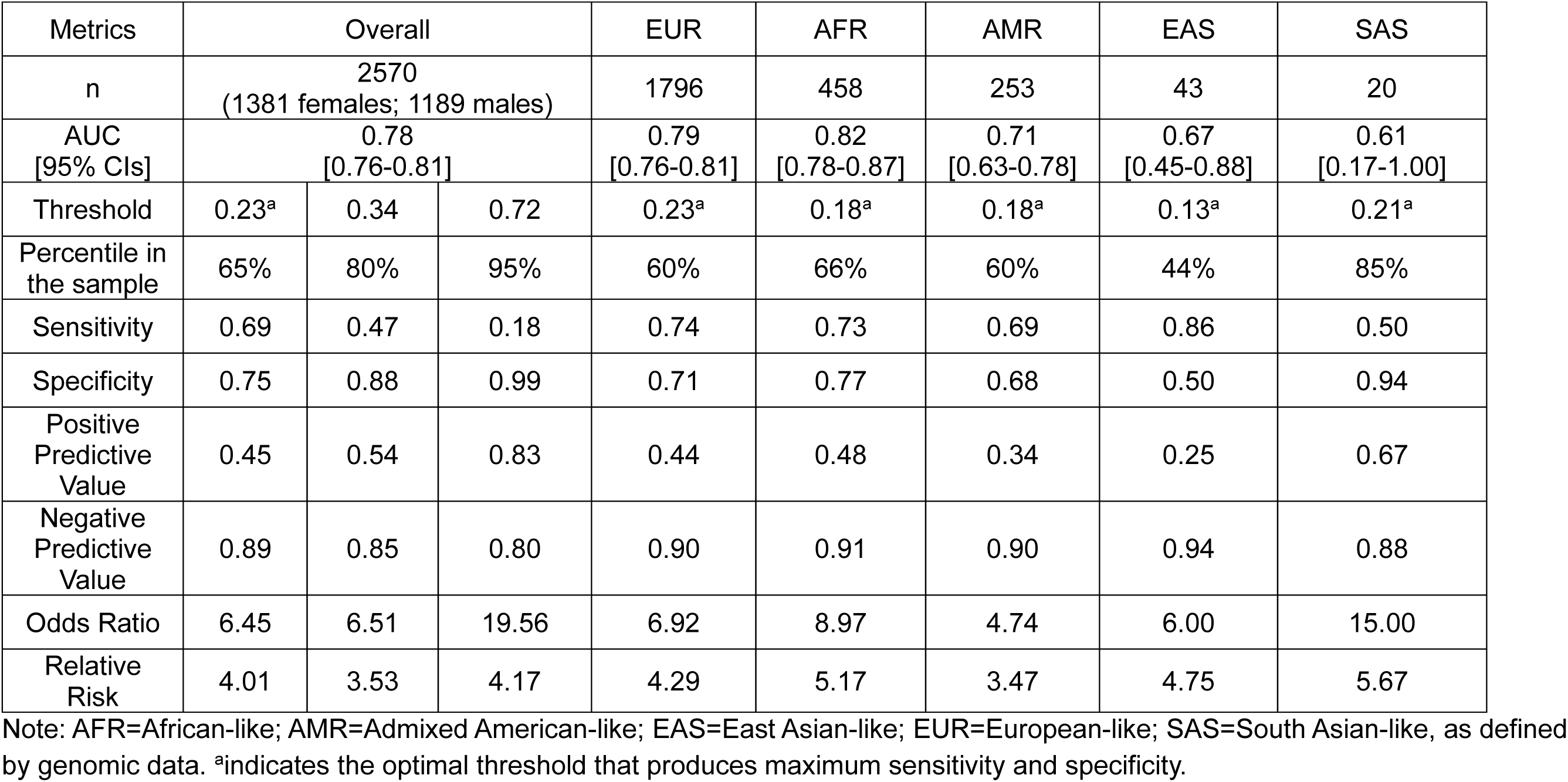
Performance Metrics for the SUD Risk Prediction Model.

Calibration analyses indicated good agreement between predicted and observed risk. The calibration plot (Figure 1) showed slight underestimation at lower predicted risk and modest overestimation at higher predicted risk.

Table 3 presents estimated ORs for each of the predictors. Male sex was associated with higher risk of any SUD (OR=1.62 [1.45-1.81]). Among genetic predictors, only the EXT PGS was significantly associated with any SUD (OR=1.15 [1.08-1.22]). All BERI items were significantly associated with SUD risk, except for trauma/stressful experiences.

**Table 3.** Estimated Odds Ratios [and 95% Confidence Intervals] for Predictors Across Outcomes.

|  | Any SUD | AUD | TUD | CUD |
| --- | --- | --- | --- | --- |
| Sex (Female as reference) | **1.62**<br>[1.45–1.81] | **1.53**<br>[1.35–1.74] | **1.48**<br>[1.23–1.78] | **1.89**<br>[1.59–2.26] |
| EXT PGS | **1.15**<br>[1.08–1.22] | **1.08**<br>[1.01–1.16] | **1.24**<br>[1.12–1.37] | **1.13**<br>[1.03–1.24] |
| PAU-specific PGS | 1.03<br>[0.98–1.09] | **1.08**<br>[1.01–1.15] | 0.90<br>[0.82–0.99] | 0.95<br>[0.88–1.03] |
| CUD-specific PGS | 1.00<br>[0.95–1.05] | 0.97<br>[0.91–1.03] | 1.01<br>[0.92–1.11] | 1.05<br>[0.97–1.14] |
| OUD-specific PGS | 1.05<br>[0.99–1.10] | 1.00<br>[0.94–1.06] | 1.00<br>[0.92–1.09] | 1.06<br>[0.98–1.15] |
| PTU-specific PGS | 1.03<br>[0.98–1.09] | 0.98<br>[0.92–1.04] | **1.13**<br>[1.03–1.25] | 0.96<br>[0.89–1.05] |
| INT PGS | 1.04<br>[0.98–1.11] | 1.01<br>[0.95–1.08] | **1.13**<br>[1.03–1.25] | 1.02<br>[0.94–1.12] |
| Childhood INT | **1.54**<br>[1.31–1.81] | 1.17<br>[0.97–1.42] | **1.60**<br>[1.23–2.07] | **1.74**<br>[1.40–2.15] |
| Childhood EXT | **2.40**<br>[2.01–2.90] | **2.48**<br>[2.05–2.99] | **1.62**<br>[1.22–2.15] | **1.81**<br>[1.45–2.11] |
| Early initiation | **1.87**<br>[1.65–2.11] | **1.78**<br>[1.54–2.06] | **1.64**<br>[1.29–2.07] | **1.75**<br>[1.44–2.11] |
| ALC use | **1.65**<br>[1.37–1.98] | **2.45**<br>[2.04–2.93] | **1.30**<br>[1.01–1.69] | 1.23<br>[0.99–1.53] |
| TOB use | **1.69**<br>[1.44–1.99] | **1.26**<br>[1.06–1.50] | **4.13**<br>[3.31–5.17] | **1.26**<br>[1.02–1.56] |
| CAN use | **3.63**<br>[2.93–4.49] | **1.57**<br>[1.27–1.95] | **1.45**<br>[1.08–1.93] | **6.69**<br>[5.33–8.41] |
| Low SES | **0.86**<br>[0.76–0.98] | **0.73**<br>[0.63–0.86] | **1.42**<br>[1.16–1.74] | **0.80**<br>[0.66–0.97] |
| Family SUD history | **1.20**<br>[1.06–1.37] | **1.21**<br>[1.04–1.40] | 1.11<br>[0.88–1.38] | 1.21<br>[0.99–1.48] |
| Peer substance use | **1.49**<br>[1.32–1.68] | **1.32**<br>[1.14–1.53] | **1.26**<br>[1.01–1.59] | **1.41**<br>[1.16–1.72] |
| Trauma/Stressful Experiences | 1.10<br>[0.98–1.24] | 1.05<br>[0.92–1.20] | 1.22<br>[0.99–1.50] | 1.04<br>[0.86–1.25] |

Note. Bold indicates significant effects (odds ratios confidence interval excludes 1). ALC use = frequent drinking in adolescence; AUD = alcohol use disorder; CAN = frequent cannabis use in adolescence; CUD = cannabis use disorder; EXT = externalizing; INT = internalizing; OUD = opioid use disorder; PAU = problematic alcohol use; PGS = polygenic scores; PTU = problematic tobacco use; TOB use = frequent smoking in adolescence; TUD: tobacco use disorder; SES = socioeconomic status; SUD = substance use disorder.

### Model Performance for Specific SUD Risks

DeLong’s test indicated no significant difference in discrimination between the AUD-specific model and the SUD model applied to AUD (AUC_AUD_=0.74; AUC_SUD_=0.74; p=.514). In contrast, significant differences were observed for TUD (AUC_TUD_=0.84; AUC_SUD_=0.80; p<.001) and for CUD (AUC_CUD_=0.86; AUC_SUD_=0.82; p<.001), with models trained on substance-specific outcomes demonstrating superior predictive performance. Figure 2 presents the receiver operating characteristic curves for AUD, TUD, and CUD, as compared to the general model for SUD.

**Figure 2.**
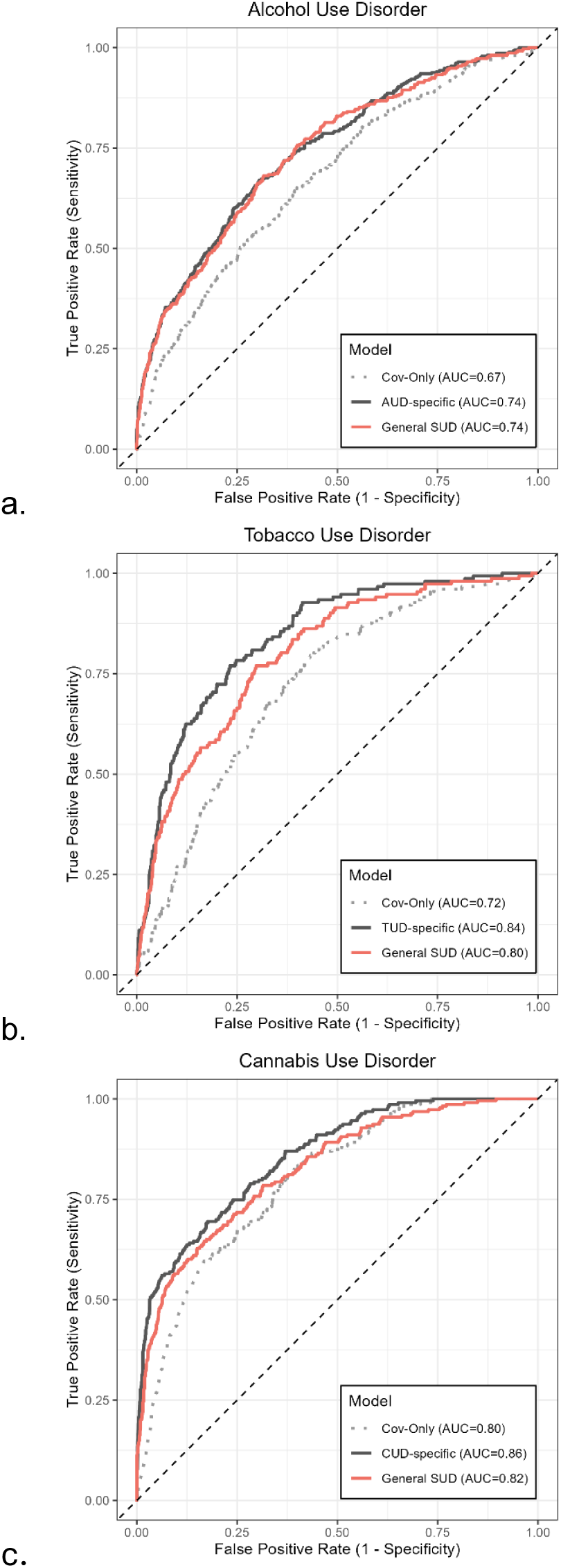
Receiver Operating Characteristic Curves of the Any SUD Model Applied to Specific SUD Outcomes Note: (a) receiver operating characteristic curve (ROC) of alcohol use disorder (AUD), (b) ROC of tobacco use disorder (TUD), (c) ROC of cannabis use disorder (CUD). Cov = covariates (sex, age of assessment, and cohort).

Table 3 includes the ORs for each of the predictors for the substance-specific models, as compared to the SUD model. The ORs indicate that the differences between models largely reflect that the frequent use of a particular substance during adolescence, and the substance specific PGS, were most predictive of the corresponding disorder (e.g., frequent use of tobacco and the problematic tobacco use specific PGS are most strongly related to TUD).

### Model Performance Across Sex

Sex-stratified prediction analyses were conducted for models for each SUD. Statistically significant sex differences in model performance were observed for SUD (AUC_Male_=0.74; AUC_Female_=0.79; p=.032). There was a significant interaction between early substance use initiation and sex (β=0.48, p=.002), whereby the association was stronger in females (OR=2.43, 95% CI [2.03, 2.91]) than in males (OR=1.51, 95% CI [1.27, 1.78]). However, including this interaction term did not improve model discrimination (AUC=0.78 [0.76–0.80], compared to AUC=0.78 from model without interaction term).

No statistically significant differences in predictive performance between males and females were found for the AUD model (AUC_Male_=0.72; AUC_Female_=0.75; p=.211), for the TUD model (AUC_Male_=0.82; AUC_Female_=0.85; p=.466), or for the CUD model (AUC_Male_=0.82; AUC_Female_=0.87; p=.097).

### Model Performance Across Genetic Similarity Groups

Model performance of the general SUD model across genetic similarity groups in the validation sample is summarized in Table 2. The model performed comparably across groups, particularly among participants of European-like, African-like, and admixed American-like genetic similarity groups. AUC estimates were less precise, with larger 95% confidence intervals, in East Asian– like and South Asian–like groups, reflecting considerably smaller sample sizes.

## Discussion

There is an unmet need to develop clinically actionable prediction models that allow individuals and clinicians to understand risk for developing a SUD before problems emerge. Our study aimed to address that gap by leveraging genetic, behavioral, and environmental information from multiple, longitudinal samples to develop and evaluate clinical prediction models for SUD risk. Our analyses build upon decades of research on the developmental nature of SUDs,^11–17,24^ coupled with recent advances in gene identification.^19,23,42^ We find that a multivariable model demonstrates strong predictive performance. The model identified approximately two-thirds of individuals who subsequently developed a SUD. Because PPV depends on the prevalence of SUD in the population studied, this metric should be interpreted relative to our sample. PPV was approximately two-fold higher than SUD prevalence (17% from the U.S. national survey on drug use^2^ and 22% in the current sample), indicating the model can meaningfully identify a group of people who are at elevated risk for developing a SUD. As such, its integration with established prevention and early intervention strategies may offer an opportunity to reduce downstream morbidity and mortality, including overdose and SUD-related subsequent medical conditions, as well as the myriad of individual, familial, and social costs associated with SUDs.

Current substance use screening measures are designed to detect substance use and symptoms. At this time, the only information that is used clinically (albeit rarely^5–7^) to indicate potential risk of developing a future SUD is family history.^43,44^ Prior studies suggest a 2-3-fold increased risk among those with a positive history.^45^ In our sample, a model including only family history similarly yielded a 3.17-fold increased risk of SUD. When family history alone was used as a predictor, model discrimination was modest (sensitivity, 0.64; specificity, 0.65; PPV, 0.35). In contrast, our full model, incorporating genetic predictors and additional BERI items, improved discrimination and risk stratification over family history alone (sensitivity, 0.69; specificity, 0.75; PPV, 0.45). Other studies have also shown the incorporation of PGS improves prediction beyond family history alone.^46^

The general SUD model also performed adequately (AUCs = 0.74-0.82) when predicting the specific common substance use disorders (AUD, TUD, CUD) evaluated here. Although significant differences in model fit were observed for TUD and CUD, the differences in AUC were trivial (ΔAUC, 0.02–0.04) and largely reflect the fact that use of a specific substance was most predictive of development of problems for that substance. Similarly, although model performance was slightly lower in males than in females, the addition of interaction terms did not meaningfully improve model performance. Accordingly, we suggest that implementing the general SUD prediction algorithm would be a parsimonious approach that has the added advantage of raising awareness about how risk for SUD is shared across substances. This may help providers and patients recognize that elevated risk for one SUD can also reflect broader vulnerability to other substance-related problems if other substances are to be used regularly.

### Limitations

These findings should be interpreted in the context of the following limitations. First, although the overall sample size suggested adequate power for model development, the relatively low prevalence of TUD and CUD relative to other outcomes may have resulted in less precise estimates and greater variability in performance metrics. Relatedly, we were unable to examine model generalizability to less common specific SUDs (e.g., opioid use disorder) due to the limited number of cases in the analytic sample. Second, the PGS were derived from GWAS findings conducted predominantly in individuals of European genetic ancestry, and the predictive performance of PGS is considerably reduced when applied to other genetic-ancestral groups.^47,48^ Nonetheless, as shown in Table 2, model performance was comparable across European, African, and admixed American populations; lower performance in East and South Asian populations likely reflects small sample sizes. Ongoing efforts by large, international GWAS consortia^20,49^ to include more diverse discovery samples are expected to improve this limitation in the future. In addition, some of the PGS (e.g., CUD-specific PGS) used in the current model may be underpowered,^19^ and continued advances in GWAS are likely to further enhance predictive performance. Importantly, PGS for SUD are probabilistic, not deterministic. Providing personalized feedback in ways that carefully consider ethical concerns and weigh the potential for benefit versus harm will be paramount when applying these findings in clinical settings.^50–53^ Lastly, although the model was evaluated using held-out testing/validation data, these data came from the same cohorts as the training data. Future validation in fully independent, external samples will be important to confirm generalizability.

## Conclusion

Tools that enable clinicians and patients to understand risk for SUD may provide an effective avenue to reduce the substantial personal and societal burden associated with SUDs. Over 80% of individuals report wanting access to personalized risk information for substance use outcomes,^54^ and initial studies by our team indicate high satisfaction with the provision of personalized feedback about substance use risk profiles.^51^ This study moves us closer to that goal by developing and evaluating a prediction model for SUD. Our findings provide a basis for future studies evaluating the implementation of such tools to guide targeted prevention efforts.

Conflicts of Interest and Financial Disclosures:

Danielle Dick is a Co-founder and Chief Scientific Officer for Thrive Genetics, Inc. She is on the Advisory Board for the Seek Women’s Health Company and HumanUp. She has received royalties from Penguin Random House for her book, The Child Code: Understanding Your Child’s Unique Nature for Happier, More Effective Parenting. The rest of the authors have no direct or indirect interests related to the work submitted for publication.

## Funding

Research reported in this publication was supported by the National Institute on Alcohol Abuse and Alcoholism and the National Institute of Drug Abuse of the National Institutes of Health under award numbers U10AA008401, P50AA022537, R01DA050721, R01AA015416, T32AA032229 and T32DA55569

## Data availability

All data sources are described in the manuscript and supplemental information. No new data were collected. Only data from existing studies or study cohorts were analyzed, some of which have restricted access to protect the privacy of the study participants. Add Health genetic data obtained through dbGaP (Study Accession: phs001367.v1.p1). Instructions on gaining access to Add Health restricted use data can be found at: https://data.cpc.unc.edu/projects/2/view. COGA genetic data available through dbGaP (Study Accession: phs000763.v1.p1). The informed consent obtained from ALSPAC (Avon Longitudinal Study of Parents and Children) participants does not allow the data to be made available through any third party maintained public repository. Supporting data are available from ALSPAC on request under the approved proposal number, B3933. Full instructions for applying for data access can be found here: http://www.bristol.ac.uk/alspac/researchers/access/. The ALSPAC study website contains details of all available data (http://www.bristol.ac.uk/alspac/researchers/our-data/). The process for obtaining the GWAS summary statistics used in these analyses are described in the corresponding original GWAS publications.

## Acknowledgement

We are extremely grateful to all the families who took part in this study, the midwives for their help in recruiting them, and the whole ALSPAC team, which includes data collection staff, data and administrations staff, technical managers and the technical staff with the Bristol Bioresource Laboratory, based within the University of Bristol. The UK Medical Research Council and Wellcome (Grant ref: MR/Z505924/1) and the University of Bristol provide core support for ALSPAC. A comprehensive list of grants funding is available on the ALSPAC website (http://www.bristol.ac.uk/alspac/external/documents/grant-acknowledgements.pdf). The Collaborative Study on the Genetics of Alcoholism (COGA), Principal Investigators B. Porjesz, V. Hesselbrock, A. Agrawal; Scientific Director, A. Agrawal; Translational Director, D. Dick, includes ten different centers: University of Connecticut (V. Hesselbrock); Indiana University (H.J. Edenberg, T. Foroud, Y. Liu, M.H. Plawecki); University of Iowa Carver College of Medicine (S. Kuperman, A. Anderson); SUNY Downstate Health Sciences University (B. Porjesz, J. Meyers); Washington University in St. Louis (L. Bierut, A. Agrawal, S. Hartz); University of California at San Diego (M. Schuckit); Rutgers University (D. Dick, R. Hart, J. Salvatore, J. Tischfield); The Children’s Hospital of Philadelphia, University of Pennsylvania (L. Almasy); Icahn School of Medicine at Mount Sinai (A. Goate, P. Slesinger); and Howard University (D. Scott). Other COGA collaborators include: C. Holzhauer, M. Hesselbrock (University of Connecticut); D. Lai, J. Nurnberger Jr., L. Wetherill, A. Miller, X. Xuei, S. O’Connor, (Indiana University); J. Kramer (University of Iowa), G. Chan (University of Iowa; University of Connecticut); C. Kamarajan, A. Pandey, D.B. Chorlian, P. Barr, S. Kinreich, G. Pandey, Z. Neale, C. Chatzinakos, J. Zhang, S. Saenz deViteri, R. Christian, A. Bingly (SUNY Downstate); G. Pathak (Icahn School of Medicine at Mount Sinai); A. Anokhin, K. Bucholz, F. Dong, A. Hatoum, E. Johnson, V. McCutcheon, J. Rice, S. Saccone (Washington University); F. Aliev, Z. Pang, S. Kuo, S. Brislin, J. Moore (Rutgers University); A. Merikangas (The Children’s Hospital of Philadelphia and University of Pennsylvania); M. Gitik, NIAAA Staff Collaborator. We continue to be inspired by our memories of Henri Begleiter and Theodore Reich, founding PI and Co-PI of COGA, and also owe a debt of gratitude to other past organizers of COGA, including Ting- Kai Li, P. Michael Conneally, Raymond Crowe, and Wendy Reich, for their critical contributions. This national collaborative study is supported by NIH Grant U10AA008401 from the National Institute on Alcohol Abuse and Alcoholism (NIAAA) and the National Institute on Drug Abuse (NIDA). The FinnTwin12 data were supported by the National Institute on Alcohol Abuse and Alcoholism of the National Institutes of Health under award numbers R01AA015416, R01AA09203, K02AA018755, and K01AA024152; and by the Academy of Finland (grants 100499, 205585,118555, 141054, 265240, 263278, and 264146). JK was supported by the Academy of Finland (grants 265240, 263278, 308248, 312073 and 352792). SA was supported by the Research Council of Finland (grant 372025 to Anu Raevuori).

## Notes

### Author Declarations

This research was reviewed and approved the Institutional Review Board at Rutgers University (Pro2022000138).

