## Supplementary for "Building a Prediction Model to Detect Elevated Risk for Developing a Substance Use Disorder"

eMethods 1

SUD Definition

SUD outcomes were harmonized to align with DSM-5 criteria, which comprise 11 symptoms spanning impaired control, social problems, risky use, and physical dependence. When craving was not assessed, symptom counts were prorated to account for the missing item. DSM-5 defines a spectrum of severity: mild (2–3 symptoms), moderate (4–5 symptoms), and severe (6 or more symptoms). In this project, we defined SUD cases as those meeting criteria for moderate or severe SUD (i.e., 4 or more symptoms), as prior research suggests that the 2-symptom threshold for mild SUD may be too lenient, raising concerns about over-diagnosis (Martin et al., 2011). Consistent with this, in the COGA dataset we found that individuals meeting criteria for mild SUD before age 25 were more likely to remain at or regress to fewer symptoms after age 25 than those meeting criteria for moderate SUD (78% vs. 47%). Participants meeting diagnostic criteria for any assessed substance were coded as cases.

Add Health. SUD was assessed by self-report questionnaire at Wave IV, when participants were approximately 24-32 years old. The questionnaire covered alcohol, tobacco, cannabis, and other self-defined favorite drugs, using items that mirror DSM-IV symptoms. The Fagerström Test for Nicotine Dependence (FTND; with a cutoff of ≥7 was used to define cases) was also administered for tobacco use. Because craving was not assessed, the maximum symptom count was 10. We prorated scores to account for the missing craving item and used the prorated score to define cases and controls.

ALSPAC. SUD was assessed by self-report questionnaire at age 24 for alcohol, tobacco, and cannabis. Symptom counts based on DSM-5 criteria were available for alcohol use. Tobacco use was assessed with the FTND. Cannabis use was assessed with the Cannabis Abuse Screening Test (CAST; Legleye et al., 2007), and a score of ≥4 was coded as indicative of problematic use.

COGA and FT12. Both cohorts used the Semi-Structured Assessment for the Genetics of Alcoholism (SSAGA), a comprehensive psychiatric interview. It assessed SUD for alcohol, tobacco, cannabis, opioids, stimulants, sedatives, hallucinogens, inhalants, and cocaine, and its items map onto all 11 DSM-5 symptoms. The FTND was also administered in both cohorts. For COGA, we used SSAGA data from annual assessments completed at age 18 or older. For FT12, we used data collected at age 22.

eMethods 2

Sensitivity Analysis

We conducted a sensitivity analysis to evaluate model performance when defining any SUD cases as meeting the threshold for mild SUD (i.e., ≥2 symptoms and/or FTND ≥5). In our sample, 34.9% of participants met this criterion.

Training our full model using this "any SUD" definition, we found that the associations between genetic, behavioral, and environmental predictors and any SUD were comparable to our main analysis, which examined any SUD defined as meeting the moderate-or-above threshold. eTable 3 presents the associations of each predictor with the mild-or-above outcome; as shown, the 95% CIs of these estimates overlapped with results from our main analysis. The primary differences were that trauma/stressful experiences were significantly associated with the mild-or-above outcome, whereas associations with low childhood socioeconomic status were attenuated.

eTable 4 summarizes model performance. Overall, predictive performance was highly consistent across the two outcome definitions. The baseline AUROC was nearly identical, and the full model showed a similar improvement over the baseline, with an AUROC increase of 0.10 in both analyses. At the optimal classification threshold, the model predicting moderate-or-above SUD demonstrated higher sensitivity (0.69) than the model predicting mild-or-above SUD (0.67). Specificity was comparable between the two models (0.75 and 0.74, respectively), indicating no meaningful sensitivity–specificity trade-off between thresholds. The positive predictive value was higher for the mild-or-above outcome, likely reflecting the higher prevalence of SUD under this broader definition. Conversely, the negative predictive value was lower for the mild-or-above outcome, which is also expected given its higher prevalence. Together, these findings indicate that the prediction model is robust to alternative case definitions and maintains similar discriminatory performance across clinically relevant SUD severity thresholds.

eMethods 3

Downsampled Summary Statistics

We recomputed the Externalizing and residual SUD summary statistics on which the polygenic scores (PGS) are based (Poore et al., 2026), removing samples that overlapped with our target sample. Updates were made to the summary statistics for lifetime cannabis use, problematic alcohol use (PAU), smoking initiation, and problematic tobacco use (PTU). For lifetime cannabis use, we used a version of the Bright (2025) lifetime cannabis use GWAS which included the All of Us and UK Biobank cohorts and excluded the ICC cohort. This version of lifetime cannabis use was correlated with at 1.03 (0.06) with the original cannabis initiation indicator. We used a version of the Zhou (2023) PAU GWAS that included MVP and UK Biobank cohorts and excluded the PGC cohorts. This version of PAU was genetically correlated at 0.96 (0.05) with the original PAU indicator. For smoking initiation, we used the Saunders (2022) smoking initiation GWAS with ALSPAC, FinnTwin12, and 23andMe removed. This version of smoking initiation was genetically correlated at 0.96 (0.03) with the original indicator, which came from the full Lui (2019) smoking initiation GWAS. Finally, the original PTU indicator was a meta-analysis of the Hancock (2018) FTND GWAS and Liu (2019) cigarettes per day (CPD). For the purposes of these analyses, we meta-analyzed FTND with CPD from the Saunders (2022) GWAS with ALSPAC, FinnTwin12, and 23andMe removed. The genetic correlation between this downsampled meta-analysis of PTU and the original PTU indicator was 1.01 (0.05).

To confirm that the downsampled summary statistics adequately represented the original indicator, we estimated genetic correlations between the original and downsampled GWAS. Using the downsampled summary statistics, we also replicated the original Externalizing model (Poore et al., 2026). Model fit was comparable (Chisq = 982, AIC = 1024, CFI = .90, SRMR = .09) and loadings were of similar magnitude (see eTable 2). Genetic correlations between the original externalizing and affected residual SUDs were high (ranging from .98 to 1.0).

eTable 1. AUC Comparison Across Different Algorithms

| | Any SUD | AUD | TUD | CUD |
|---|---|---|---|---|
| Baseline Model | 0.68 (0.66-0.71) | 0.67 (0.64-0.70) | 0.72 (0.69-0.76) | 0.80 (0.77-0.82) |
| Full Model | | | | |
| Traditional Regression | 0.78 (0.76–0.81) | 0.74 (0.72–0.77) | 0.84 (0.81–0.87) | 0.86 (0.83–0.88) |
| LASSO | 0.78 (0.76–0.81) | 0.74 (0.71–0.77) | 0.84 (0.81–0.87) | 0.86 (0.83–0.88) |
| Ridge | 0.79 (0.76–0.81) | 0.74 (0.71–0.77) | 0.84 (0.82–0.87) | 0.85 (0.83–0.88) |

Note: The baseline model included sex, age at assessment, cohort, and genetic ancestry group. The full model additionally included genetic predictors and 10 BERI items.

eTable 2 Loadings From the Externalizing Genome-Wide Association Analysis Before and After Downsampling

| Indicator | Original loading (SE) | New loading (SE) |
|---|---|---|
| SMOK | .76 (.02) | .79 (.02) |
| ASEX | .79 (.02) | .76 (.02) |
| NSEX | .73 (.02) | .72 (.02) |
| RISK | .52 (.03) | .52 (.03) |
| CANN | .66 (.03) | .62 (.03) |
| ADHD | .65 (.04) | .66 (.04) |
| PAU | .58 (.02) | .64 (.02) |
| OUD | .79 (.03) | .80 (.03) |
| PTU | .34 (.03) | .35 (.02) |
| CUD | .85 (.02) | .86 (.03) |

Note: SMOK = smoking initiation; ASEX = age at first sexual intercourse; NSEX = number of sexual partners; RISK = general risk tolerance; CANN = cannabis initiation; ADHD = attention-deficit/hyperactivity disorder; PAU = problematic alcohol use; OUD = opioid use disorder; PTU = problematic tobacco use; CUD = cannabis use disorder.

eTable 3 Odds Ratios and 95% CIs for Predictors Across Outcomes Defined as Mild-or-Above SUD

| | **Any SUD defined by mild and above** |
|---|---|
| Sex (Female as reference) | **1.68 [1.52-1.85]** |
| EXT PGS | **1.20 [1.13-1.26]** |
| PAU-specific PGS | 1.04 [0.99-1.09] |
| CUD-specific PGS | 0.99 [0.94-1.03] |
| OUD-specific PGS | 1.03 [0.98-1.08] |
| PTU-specific PGS | 1.04 [0.99-1.09] |
| INT PGS | 1.05 [0.99-1.10] |
| Childhood INT | **1.41 [1.20-1.64]** |
| Childhood EXT | **1.96 [1.61-2.39]** |
| Early initiation | **2.11 [1.89-2.36]** |
| ALC use | **1.74 [1.43-2.12]** |
| TOB use | **2.30 [1.95-2.72]** |
| CAN use | **3.93 [3.01-5.12]** |
| Low SES | 0.92 [0.82-1.04] |
| Family SUD history | **1.28 [1.14-1.44]** |
| Peer substance use | **1.55 [1.39-1.73]** |
| Trauma/Stressful Experiences | **1.17 [1.06-1.30]** |

Note. ALC use = frequent drinking in adolescence; CAN = frequent cannabis use in adolescence; EXT = externalizing; INT = internalizing; PGS = polygenic scores; TOB use = frequent smoking in adolescence.

eTable 4 Performance Metrics for the Risk Prediction Model for Any SUD Defined as Mild or Above at the Optimal Threshold

| Metrics | Value |
|---|---|
| AUC_baseline (covariates only) | 0.68 (0.67-0.71) |
| AUC_full_model (covariates+PGS+BERI) | 0.78 (0.77-0.80) |
| Sensitivity | 0.67 |
| Specificity | 0.74 |
| Positive Predictive Value | 0.61 |
| Negative Predictive Value | 0.79 |
| Odds Ratio | 6.02 |
| Relative Risk | 2.93 |

Note: AUC = area under the receiver operating characteristic curve eFigure 1. Correlations among BERI Items.

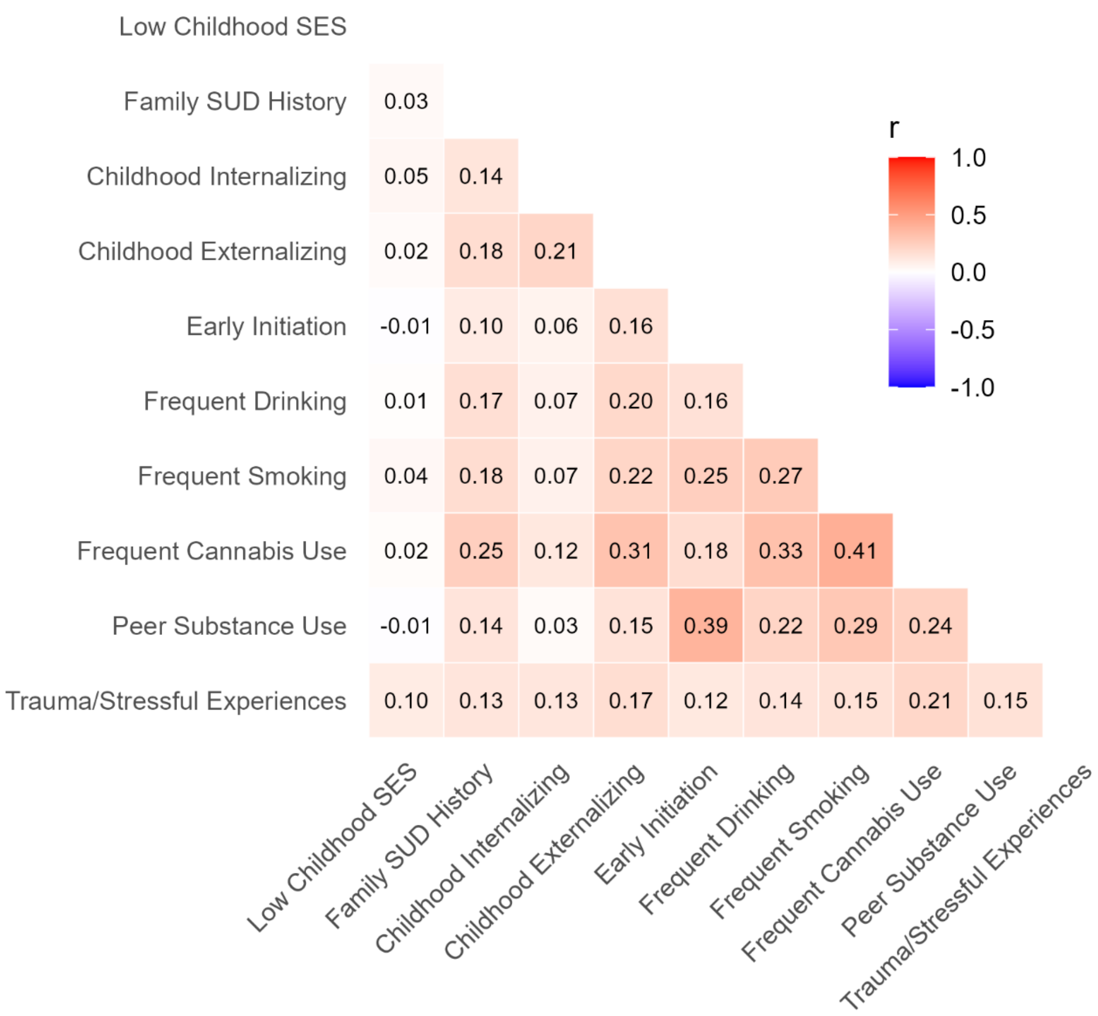


Note: SES = socioeconomic status; SUD = substance use disorder; r = tetrachoric correlation.
